# AKI Risk Score (AKI-RiSc): Developing an Interpretable Clinical Score for Early Identification of Acute Kidney Injury for Patients Presenting to the Emergency Department

**DOI:** 10.1101/2021.06.18.21259146

**Authors:** Yukai Ang, Marcus Eng Hock Ong, Feng Xie, Su Hooi Teo, Lina Choong, Riece Koniman, Bibhas Chakraborty, Andrew Fu Wah Ho, Nan Liu

## Abstract

**Background:** Acute kidney injury (AKI) in hospitalised patients is a common syndrome associated with poorer patient outcomes. Clinical risk scores can be used for the early identification of patients at risk of AKI.

**Methods:** We conducted a retrospective study using electronic health records of Singapore General Hospital emergency department patients who were admitted from 2008 to 2016. The primary outcome was inpatient AKI of any stage within 7 days of admission based on Kidney Disease Improving Global Outcome (KDIGO) 2012 guidelines. AutoScore, a machine learning based algorithm, was used to generate point based clinical scores from the study sample which was divided into training, validation and testing cohorts. Model performance was evaluated using area under the curve (AUC).

**Results:** Among the 119,468 admissions, 10,693 (9.0%) developed AKI. 8,491 were stage 1 (79.4%), 906 stage 2 (8.5%) and 1,296 stage 3 (12.1%). The AKI Risk Score (AKI-RiSc) was a summation of the integer scores of 6 variables: serum creatinine, serum bicarbonate, pulse, systolic blood pressure, and diastolic blood pressure. AUC of AKI-RiSc was 0.730 (95% CI: 0.713 – 0.747), outperforming an existing AKI Prediction Score model which achieved AUC of 0.665 (95% CI: 0.646 – 0.679) when evaluated on the same test cohort. At a cut-off of 4 points, AKI-RiSc had a sensitivity of 82.5% and specificity of 46.7%.

**Conclusion:** AKI-RiSc is a simple point based clinical score that can be easily implemented on the ground for early identification of AKI in high-risk patients and potentially be applied in healthcare settings internationally.

## Introduction

Acute kidney injury (AKI) is a common clinical syndrome affecting 10% to 20% percent of hospitalized patients worldwide.^1^ It is independently associated with increased risk of inpatient mortality,^2^ significant morbidity upon discharge,^3^ and increased healthcare costs as well as length of hospital stay.^4^ AKI is also often clinically silent and may not be promptly recognised by the attending physician.^5^ Established kidney injury is often difficult to treat,^6^ hence there is a need for early detection to initiate treatment promptly.

AKI is typically diagnosed by the magnitude of serum creatinine (SCr) rise and is usually based on well accepted criterion such as the Kidney Disease Improving Global Outcome AKI (2012) guidelines. Early warning systems have been developed to flag patients with pathological rises in SCr, which can then be paired with intervention care bundles that can prevent the progression of AKI. ^7^ However, increase in SCr levels can lag kidney injury by up to 48 hours.^8^ Physicians, therefore, are only reacting to damage that has already been done.

AKI prediction models are potential solutions to this problem. In recent years, there has been increasing interest in the development of machine learning (ML) models, which can accurately predict AKI development in patients before any rise in SCr.^9–11^ Physicians will then be able to intervene earlier and halt the progression of the renal insult.^12^ However, these ML models have yet to be widely utilized in clinical practice due to their complexity and difficulty of implementation in existing hospital IT systems.^11^ Furthermore, many ML models derive their predictions via black box approaches, which are not always be explainable to humans in a rational way. Hence, clinicians may not be as inclined to apply them in clinical practice.^13^ Point based prediction scores offer a simple and interpretable solution to these ML models. However, most existing point based AKI risk scores were designed for specific patient populations in the Intensive Care Unit,^14,15^ post-operative period,^16^ and undergoing procedures involving contrast,^16–18^ when the majority of AKI occurs in the general ward setting.^19^

The primary aim of this study was to create a simple point based clinical AKI Risk Score (AKI-RiSc) for the general patient population using a systematic, machine learning-based scoring framework – AutoScore.^20^ The AKI-RiSc was designed to assess a patient’s 7-day inpatient risk of AKI development in the setting of the emergency department (ED). It is envisioned that AKI-RiSc can function as a convenient and informative adjunct in allowing clinicians to more accurately assess a patient’s risk of inpatient AKI and institute any necessary management steps.

## Methods

### Study design and setting

We conducted a retrospective, single-centre study in Singapore General Hospital (SGH) to derive AKI-RiSc using EHR of the patients in the ED and inpatient wards. SGH is a tertiary hospital that receives over 120,000 ED visits and 36,000 admissions annually. A waiver of consent for EHR data collection was granted, and the study protocol was approved by Singapore Health Services’ Centralised Institutional Review Board.

### Study sample

All adult patients (>18 years old) visiting the ED between 1 January 2008 to 31 December 2016 and who were subsequently admitted to medical and surgical wards were studied.^21^ Patients were excluded if they met any of the following criteria: 1) patients with no records of SCr, 2) patients with AKI on presentation at the ED, defined based on KDIGO change of SCr from median annualised SCr baseline, 3) patients with pre-existing advanced chronic kidney disease (CKD) based on KDIGO guidelines (SCr ≥353.6 µmol/L on admission) and 4) patients with no information of comorbidities. Patients were then followed up to 7 days post-admission to determine if they developed AKI or not.

### AKI definition and outcomes

The primary outcome of this study was the development of inpatient AKI of any stage within 7 days of admission from the ED. We used the National Health Service (NHS) automated AKI algorithm based on KDIGO guidelines to define AKI.^22^ The algorithm had been applied in clinical practice and was chosen because of its ability to account for patients with and without prior baseline SCr information.^23^ A patient was determined to have AKI if any of the three criteria were met: 1) Increase in SCr to ≥1.5 times of the median of all SCr readings 8 to 365 days ago, 2) increase in SCr to ≥1.5 times of the lowest SCr reading in the past 7 days, 3) increase in SCr by ≥ 26.5µmol/L within 48 hours from the lowest SCr reading. Other outcomes that were examined in this study were patients who developed at least Stage 2 AKI and patients who developed Stage 3 AKI.

### Data collection and candidate variables

We extracted the data from the hospital’s EHR – SingHealth Electronic Health Intelligence System (eHints). The data was de-identified, and the death records were obtained from the national death registry and matched to specific patients in the EHR. As the AKI-RiSc was designed to be applied in the ED setting, we only selected variables that were exclusively available and reliably obtained at the ED. The final 33 candidate variables were selected based on literature review and expert opinion from clinicians, which consisted of patient demographics, vital signs, biochemical results, comorbidities, medical interventions, and visits to hospital – as seen in Table 1. Comorbidities were obtained from the hospital diagnosis and discharge records in the past five years before each patient’s current ED visits. They were recorded as the International Classification of Diseases (ICD) 9/10 codes.^24^

**Table 1:**
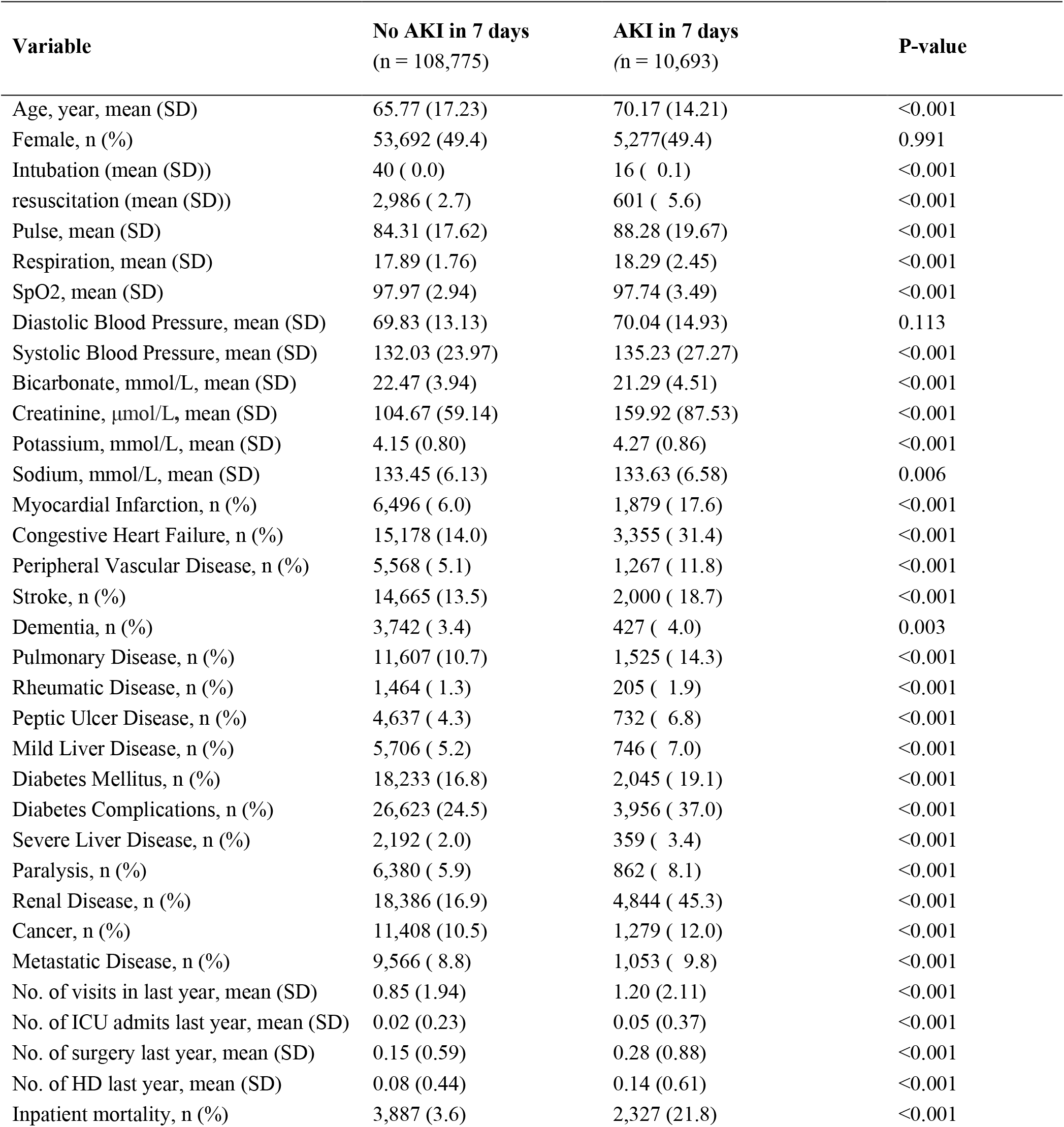
Comparison of patient characteristics between those who developed AKI 7 days after admission and those who did not at the emergency department.

### Statistical analyses and predictive modeling

Variables were included for the analysis only if more than 80% of them were available in the study cohort. Chi-square for categorical variables and *t-*tests for continuous variables were used where appropriate to compare between the patients who developed AKI and those who did not. Univariable analysis was also used to determine the odds ratios of risk factors between the two groups. Figure 1 depicts data preparation. Patients admitted between 2008 to 2015 were randomly divided into derivation and validation cohorts, at a 70% and 30% ratio, respectively. (Figure 1b) Patients admitted in 2016 were used as the test cohort to evaluate the final score.

**Figure 1:**
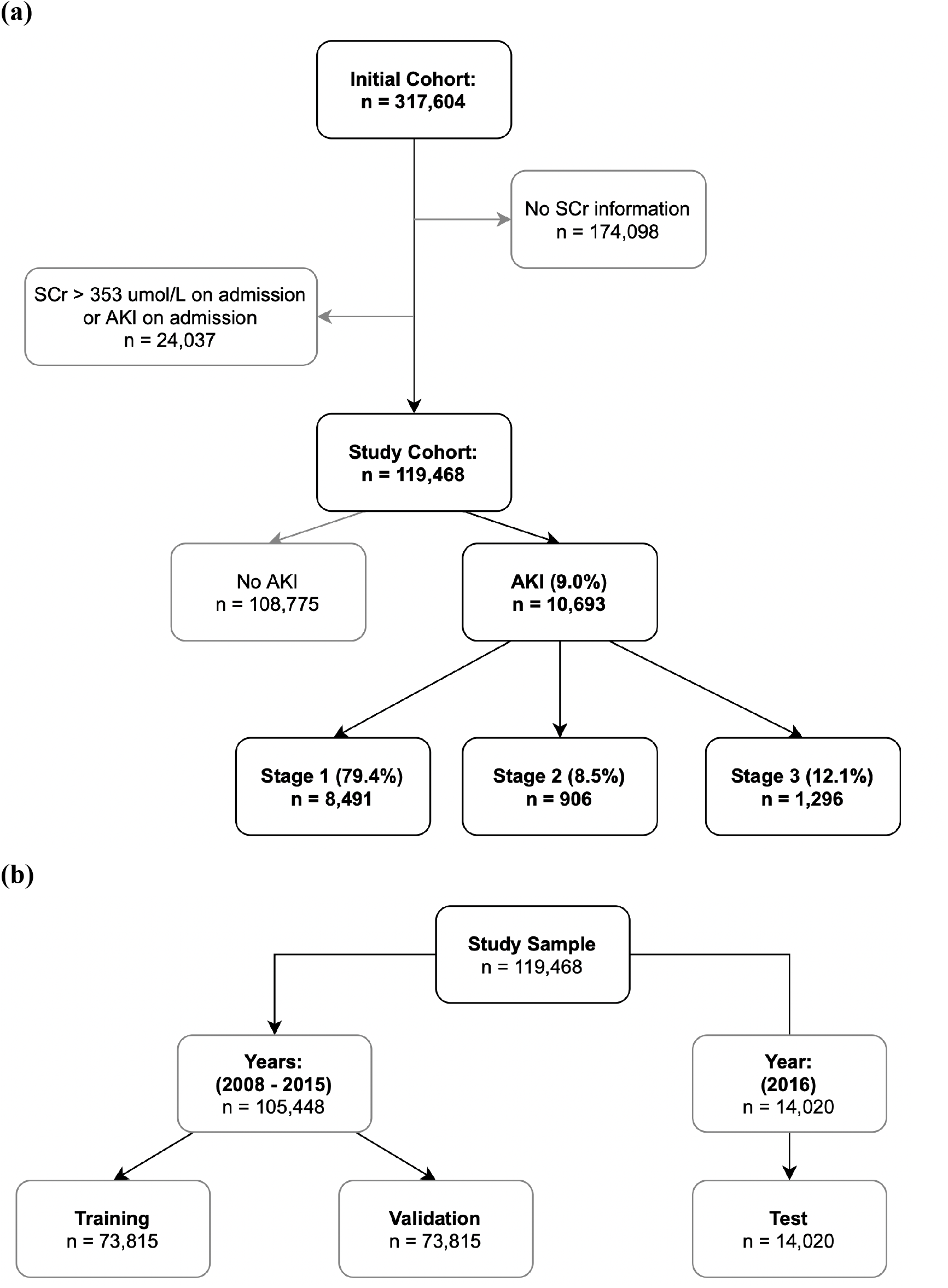
Flowcharts of (a) patient selection, and (b) splitting of study population into training, validation and test cohorts

AKI-RiSc was developed using AutoScore,^20^ a machine learning-based algorithm for interpretable clinical score generation. AutoScore allows for the easy and transparent development of interpretable clinical score models for a pre-defined clinical outcome, using a combination of machine learning, logistic regression, and user-defined parameter fine-tuning. Figure S1 illustrates the process of score derivation and validation. Details about AutoScore framework can be found under the eMethods in the supplementary paper.

The training cohort was first used to generate the preliminary AKI-RiSc models using the AutoScore main flow. The validation cohort was then used to evaluate the performance of various candidate AKI-RiSc models and allow for parameter fine-tuning. After selecting the final AKI-RiSc model, we evaluated its performance using the testing cohort, and bootstrapped samples were applied to calculate 95% confidence intervals. The primary outcome measure of inpatient AKI development was used for model derivation and model testing. The predictive power of AKI-RiSc was assessed using the area under the curve (AUC) in the receiver operating characteristic (ROC) analysis. Sensitivity, specificity, positive predictive value (PPV), and negative predictive value (NPV) of the various AKI-RiSc cut-offs were also calculated. We also compared AKI-RiSc with an existing AKI clinical score, the AKI Prediction Score (APS)^25^, by evaluating its performance using the same testing cohort with the same primary outcome measure.

## Results

The initial cohort comprised 317,604 unique visits to the ED that were subsequently admitted. After excluding admissions with no available SCr (n = 174,098), and those with SCr >353.6 µmol/L or AKI on admission (n = 24,037), we arrived at a final cohort of 119,468 unique entries. Among this cohort, 10,693 (9.0%) developed AKI of any stage within seven days of admission, of which 8,491 were stage 1 (79.4%), 906 stage 2 (8.5%), and 1,296 stage 3 (12.1%) (Figure 1). The overall in-hospital mortality rate of patients admitted with AKI was 21.8%, as opposed to 3.6% in non-AKI counterparts. While most AKI patients were stage 1, the number of patients in stage 3 was higher than stage 2, which was also observed in another recent hospital-wide AKI study.^26^

Patients who developed AKI were older, had higher baseline SCr, higher admission SCr, and had more comorbidities associated with them (Table 1). Univariable analysis (Table 2) identified tachycardia, tachypnoea, hypotension, and hypertension as the most significant risk factors among the vital signs. Higher SCr levels on admission at the ED also increased the risk of inpatient AKI. Among the other lab values, low serum bicarbonate was the most significant risk factor for developing AKI. Extremes of serum sodium and potassium values also conferred an increased risk of AKI. All comorbidities were significantly associated with the development of AKI, but patients with renal disease had the greatest risk (odds ratio [OR]: 4.07, 95% confidence interval [CI] 3.91 – 4.24). Previous surgeries, admissions to the hospital, high dependency unit, and ICU were also significant risk factors.

**Table 2:** Univariable analysis of candidate variables

| Variable | Odds Ratio (95% Confidence interval) | P-value |
| --- | --- | --- |
| Gender, Male | 1.00 (0.96-1.04) | 0.98 |
| Age, years (Ref: <32) |  |  |
| 32 – 52 | 2.38 (2.00-2.84) | <0.001 |
| 53 – 80 | 4.10 (3.48-4.83) | <0.001 |
| 81 – 88 | 4.66 (3.94-5.52) | <0.001 |
| >88 | 4.69 (3.92-5.61) | <0.001 |
| Pulse, beats per min (Ref: 59 – 68) |  |  |
| 31 – 58 | 1.15 (1.03-1.29) | 0.01 |
| 69 – 98 | 1.18 (1.11-1.26) | <0.001 |
| 99 – 115 | 1.59 (1.48-1.71) | <0.001 |
| >115 | 2.35 (2.15-2.57) | <0.001 |
| Respiration, breaths per min (Ref: 16) |  |  |
| <16 | 2.45 (2.14-2.8) | <0.001 |
| 17 | 1.04 (0.96-1.12) | 0.35 |
| 18 – 19 | 1.15 (1.08-1.22) | <0.001 |
| >19 | 2.23 (2.08-2.39) | <0.001 |
| SpO2, % (Ref: >96) |  |  |
| 95 – 96 | 1.19 (1.13-1.25) | <0.001 |
| <95 | 2.14 (1.92-2.38) | <0.001 |
| SBP, mmHg (Ref: 98 – 110) |  |  |
| <98 | 1.39 (1.25-1.53) | <0.001 |
| 111 – 150 | 0.96 (0.90-1.02) | 0.16 |
| 151 – 176 | 1.28 (1.2-1.38) | <0.001 |
| >177 | 1.80 (1.64-1.96) | <0.001 |
| DBP, mmHg (Ref: 59 – 79) |  |  |
| <50 | 1.79 (1.64-1.95) | <0.001 |
| 50 – 58 | 1.14 (1.08-1.21) | <0.001 |
| 80 – 92 | 1.16 (1.09-1.22) | <0.001 |
| 93 – 148 | 1.55 (1.43-1.68) | <0.001 |
| Bicarbonate, mmol/L (Ref: 25.4 – 28.4) |  |  |
| <15.8 | 3.11 (2.85-3.4) | <0.001 |
| 15.8 – 19.2 | 1.95 (1.81-2.1) | <0.001 |
| 19.3 – 25.3 | 1.18 (1.10-1.26) | <0.001 |
| >28.4 | 1.27 (1.14-1.41) | <0.001 |
| Creatinine, micromol/L (Ref: 46 – 61) |  |  |
| <46 | 2.25 (1.97-2.56) | <0.001 |
| 62 – 143 | 2.04 (1.86-2.22) | <0.001 |
| 144 – 254 | 6.11 (5.57-6.70) | <0.001 |
| 255 – 366 | 14.04 (12.71-15.51) | <0.001 |
| Potassium (Ref: 3.5 – 4.6) |  |  |
| <3 | 1.26 (1.14-1.39) | <0.001 |
| 3 – 3.4 | 0.96 (0.90-1.03) | 0.27 |
| 4.7 – 5.5 | 1.46 (1.38-1.53) | <0.001 |
| >5.5 | 1.58 (1.46-1.71) | <0.001 |
| Sodium (Ref: 129-137) |  |  |
| <122 | 1.18 (1.07-1.29) | <0.001 |
| 122 – 128 | 0.98 (0.92-1.05) | 0.62 |
| 138 – 140 | 1.03 (0.98-1.09) | 0.26 |
| >140 | 1.38 (1.28-1.48) | <0.001 |
| Renal Disease | 4.07 (3.91-4.24) | <0.001 |
| Myocardial Infarction | 3.36 (3.17-3.55) | <0.001 |
| Congestive Heart Failure | 2.82 (2.70-2.95) | <0.001 |
| Peripheral Vascular Disease | 2.49 (2.34-2.66) | <0.001 |
| Stroke | 1.48 (1.40-1.55) | <0.001 |
| Dementia | 1.17 (1.05-1.29) | <0.001 |
| Pulmonary Disease | 1.39 (1.31-1.47) | <0.001 |
| Rheumatic Disease | 1.43 (1.24-1.66) | <0.001 |
| Peptic Ulcer Disease | 1.65 (1.52-1.79) | <0.001 |
| Mild Liver Disease | 1.35 (1.25-1.47) | <0.001 |
| Severe Liver Disease | 1.69 (1.51-1.89) | <0.001 |
| Diabetes Mellitus | 1.17 (1.12-1.24) | <0.001 |
| Diabetic Complications | 1.81 (1.74-1.89) | <0.001 |
| Paralysis | 1.41 (1.31-1.52) | <0.001 |
| Cancer | 1.16 (1.09-1.23) | <0.001 |
| Metastatic Disease | 1.13 (1.06-1.21) | <0.001 |
| High dependency admissions in last year (Ref: 0) |  |  |
| 1 | 1.64 (1.47-1.82) | <0.001 |
| 2 | 1.76 (1.52-2.03) | <0.001 |
| >2 | 2.12 (1.79-2.52) | <0.001 |
| ICU admissions in last year (Ref: 0) |  |  |
| 1 | 2.11 (1.74-2.55) | <0.001 |
| 2 | 2.50 (1.96-3.17) | <0.001 |
| >2 | 2.15 (1.56-2.96) | <0.001 |
| Hospital admissions in last year (Ref: 0) |  |  |
| 1 | 1.49 (1.42-1.57) | <0.001 |
| 2 | 1.77 (1.65-1.89) | <0.001 |
| >2 | 2.02 (1.90-2.14) | <0.001 |
| Number of surgeries in last year (Ref: 0) |  |  |
| 1 | 1.44 (1.34-1.55) | <0.001 |
| 2 | 1.75 (1.56-1.96) | <0.001 |
| >2 | 2.58 (2.28-2.91) | <0.001 |
| Intubation | 4.07 (2.28-7.28) | <0.001 |
| Resuscitation | 2.11 (1.93-2.31) | <0.001 |

AKI-RiSc was developed in a stepwise manner with the AutoScore framework. After ranking the essential AKI prediction variables, a parsimony plot (Figure 2) was created to visualise model selection. The variables shown in Figure 2 were ranked in order of importance of AKI prediction based on the initial random forest selection, with SCr being the most important and intubation being the least. Figure 2 also plots the model performance (AUC) against model complexity (number of variables). The most significant increase in model performance was observed from the first to the sixth variable, with marginal gains in performance fort every other variable added thereafter.

**Figure 2:**
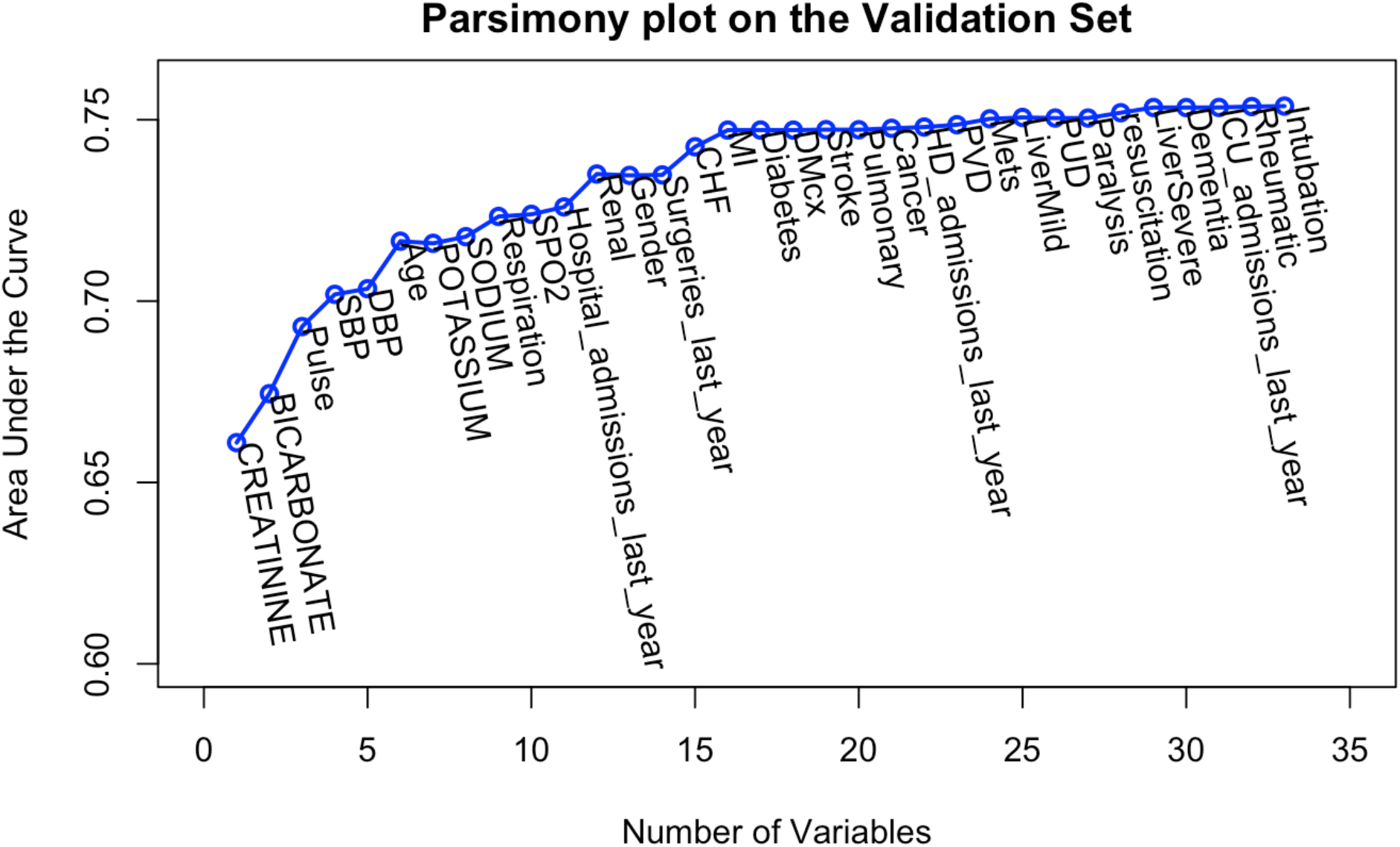
Parsimony plot of variables and predictive performance of model on the validation set.

We selected the six most important variables in the parsimony plot (SCr, serum bicarbonate, pulse, SBP, DBP, and age) to build the AKI-RiSc. Each variable was allocated an integer score based on its importance. The final score was a summation of all the variable’s assigned scores, ranging from 0 to 15 (Table 3). Risk of inpatient AKI development was positively correlated with the higher AKI-RiSc scores. Higher SCr levels contributed most significantly with greater final scores, with SCr values ≥250 μmol/L being allocated the highest score of 7. Tachycardia (≥120 beats per minute) was also given a higher score of 3. Other variables associated with higher scores were lower bicarbonate levels of <20 mmol/L, systolic blood pressure of ≥150 mmHg, diastolic blood pressure of ≥90 mmHg, and older age ≥50.

**Table 3:** Six-variable AKI Risk Score (AKI-RiSc) breakdown

| Predictor | Allocated Score |  |  |  |  |  |
| --- | --- | --- | --- | --- | --- | --- |
|  | 0 | 1 | 2 | 3 | 5 | 7 |
| Creatinine<br>($\mu\text{mol/L}$) | <100 | | 100 – 149 | | 150 – 249 | $\geq 250$ |
| Bicarbonate<br>($\mu\text{mol/L}$) | $\geq 20$ | <20 | | | | |
| Pulse<br>(beats/min) | <100 | | 100 – 119 | $\geq 120$ | | |
| SBP<br>(beats/min) | <150 | $\geq 150$ | | | | |
| DBP<br>(beats/min) | <90 | $\geq 90$ | | | | |
| Age | <50 | | $\geq 50$ | | | |

SBP: Systolic blood pressure, DBP: Diastolic blood pressure

In terms of performance, AKI-RiSc performed reasonably well at an AUC of 0.730 (95% CI: 0.713 – 0.747) when evaluated on the test cohort. At a cut-off of 4 points, it has a sensitivity of 82.5%, specificity of 46.7%, positive predictive value of 7.7%, and negative predictive value of 97.1%. When the APS, ^25^ a similar point based AKI risk score, was evaluated on the same test cohort as a basis of comparison, it scored an AUC of 0.665 (95% CI: 0.646 – 0.679). The APS consisted of age, respiratory rate, mental status (AVPU), CKD, CHF, DM, Liver Disease. Table 4 presents the score thresholds and respective sensitivity, specificity, positive predictive value, and negative predictive value. Figure 3 shows the correlation between the calculated AKI-RiSc value and the proportion of patients who developed AKI within 7 days. Almost 30% of patients with an AKI-RiSc of 9 developed AKI.

**Table 4:** Threshold scores of the predicted risk of AKI based on the ARS, including percentage of patient within score threshold, sensitivity, specificity, positive predictive value (PPV), and negative predictive value (NPV)

| Predicted Risk of AKI | Score cut-off | Percent of patients (%) | Sensitivity (95% CI) | Specificity (95% CI) | Positive Predictive Value (95% CI) | Negative Predictive Value (95% CI) |
| --- | --- | --- | --- | --- | --- | --- |
| ≥2.5% | ≥2 | 93 | 98.5% (97.8-99.2%) | 7% (6.6-7.4%) | 7.7% (7.7-7.8%) | 98.4% (97.5-99.1%) |
| ≥5% | ≥4 | 55 | 82.5% (80-85%) | 46.7% (45.9-47.6%) | 10.9% (10.6-11.3%) | 97.1% (96.7-97.5%) |
| ≥7.5% | ≥6 | 26 | 57.6% (54.6-60.6%) | 77% (76.3-77.6%) | 16.5% (15.7-17.4%) | 95.8% (95.5-96.1%) |
| ≥10% | ≥7 | 20 | 51.9% (49-55%) | 82.2% (81.5-82.8%) | 18.8% (17.7-19.8%) | 95.6% (95.3-95.8%) |
| ≥15% | ≥8 | 14 | 41.1% (38.3-44.2%) | 88.2% (87.6-88.7%) | 21.6% (20.1-23.1%) | 95% (94.7-95.2%) |
| ≥30% | ≥11 | 2 | 8.8% (7.2-10.6%) | 98.1% (97.9-98.4%) | 27.4% (22.9-32%) | 93.1% (93-93.3%) |

**Figure 3:**
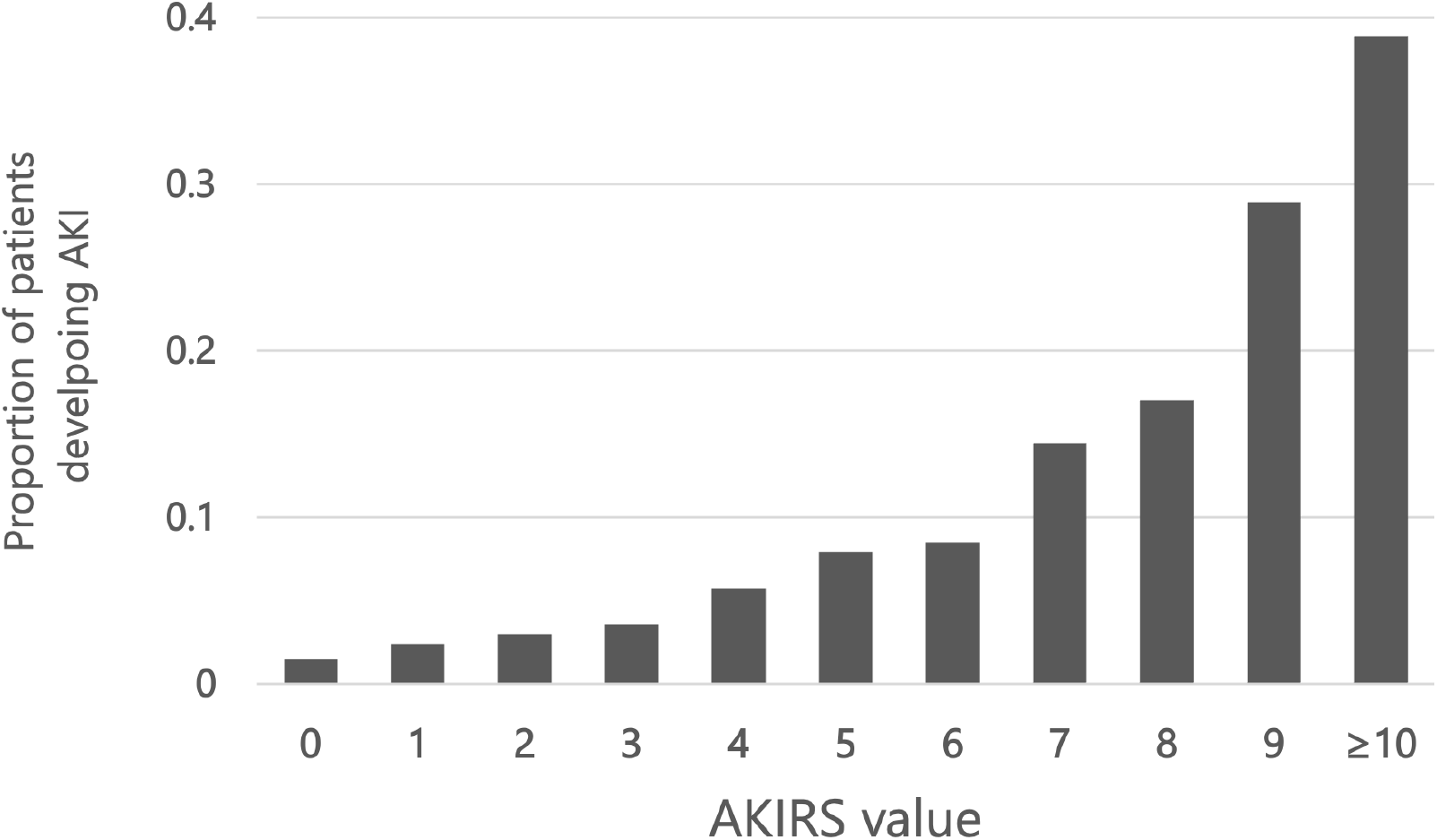
Proportion of patients developing AKI within 7 days of admission and respective AKI-RiSc value

## Discussion

In this study, we used a large retrospective dataset and the AutoScore framework to construct a point-based clinical score, AKI-RiSc, to assess the 7 day risk of AKI development in a general patient population after admission from the ED. The AKI-RiSc performed significantly better than the original APS score when evaluated on the same test cohort.

The development of AKI-RiSc using AutoScore and our dataset brings several advantages. First, our sample size of 119,468 is considerably large which improves the reliability of the results obtained from our study. Secondly, the AutoScore scoring framework is built upon machine learning, which achieves a better parsimonious solution for clinical prediction tasks with large data sets.^27^ Thirdly, the selection of variables for AKI-RiSc was also performed in a transparent manner by identifying the most parsimonious solution from the parsimony plot (Figure 2), allowing clinicians to better understand the processes involved in generating the score.

AKI-RiSc was designed for the general patient population in the form of a simple point based score that can be easily applied and interpreted by clinicians. Only the 6 most important variables were chosen for the final model, as additional variables only contributed marginal increase in model performance with the downside of increasing model complexity. (Figure 2) These variables, (SCr, bicarbonate, pulse, SBP, DBP, age) consisted of common biochemical results as well as patient parameters that could be obtained directly at the bedside within 2 hours of admission at the ED. The score’s simplicity makes it practical in the clinical setting in the form of an automated score integrated in a hospital’s her, or just via manual calculation by the clinician.^28^

When compared to the APS developed by Forni et al. in 2013, which was also a similar point-based AKI risk score for general patients in the ED,^25^ the AKI-RiSc outperformed the APS by a significant margin in terms of AUC performance (0.730 vs 0.665). One key difference between the two scores was the use of biochemical variables in AKI-RiSc – SCr and serum bicarbonate. SCr is a well-accepted marker of kidney function and has also been the most crucial variable in other AKI prediction models.^10,29^ Low bicarbonate, which typically indicates metabolic acidosis, was also found independently associated with increased AKI risk in other studies.^30^ Including these biochemical results could have potentially improved the performance of AKI-RiSc. It is worth noting that the APS also achieved a similar AUC of 0.71 when evaluated on their own UK cohort.^31^ One explanation of the poor performance of the APS in our Singapore study could be the inherent differences between the population demographics it was derived and tested on.^32^ For example, the average population age of the UK cohort was overall older than that of the Singapore cohort.

Comparing to other ML models, however, the performance of AKI-RiSc lags. Tomasev’s et al. developed a recurrent neural network AKI prediction model that calculated and updated the risk of AKI every 48 hours, performing well with an AUROC of 0.921.^29^ Despite these remarkable results, the translation of ML models to practical clinical medicine has been limited. ML models often require significant amount of information to function optimally, which may be difficult to obtain consistently.^33^ Furthermore, the complexity of the models’ algorithms make it challenging for implementation in many hospital’s IT infrastructure.^34^ Many of these machine learning models are also developed using a black box approach as opposed to more traditional logistic regression models which clinicians are familiar with.^34^ This can potentially limit data interpretation and the willingness of clinicians to adopt the model’s predicted risk.^13^ Therefore, while AKI-RiSc may not perform as well as pure ML models in AKI prediction, it has the benefit of being much more easily implemented in the clinical setting with a reasonable predictive performance.

Several actions can be taken after a high-risk patient has been identified using the AKI-RiSc in the ED. First, interventional bundle care plans can be instituted for patients identified with a high AKI-RiSc in the form of a checklist to guide clinicians in the treatment of AKI^7^ and potentially improve patient outcomes.^35^ Second, a high AKI-RiSc score can prompt the emergency physician to make earlier nephrology consults, which will improve the outcomes of patients with renal impairment.^36^ Third, the utility of the AKI-RiSc can be further enhanced when paired with novel AKI biomarkers, in which high risk patients who may benefit from AKI biomarker test can be initially identified using AKI-RiSc.

There are limitations in this study. As with most other AKI studies, AKI was defined only using serum SCr as urine output information was unavailable. Furthermore, more than half of the admissions were first-time admissions, meaning that they did not have a median annual baseline SCr. A significant number of admissions relied on the ED SCr reading as the baseline. In reality, admission SCr could already have been pathologically raised due to prior kidney insults preceding the admission. Hence, using the ED SCr as a baseline reference meant that the actual incidence of AKI could be underestimated.^37^ While we could accurately identify the diagnosis of AKI in patients by trending changes in SCr, we did not have any information on the aetiology of the AKI. However, AKI-RiSc was designed to be an alert system designed to recommend treatment, not as a diagnostic tool.^25^

The next step would be to prospectively validate the AKI-RiSc in the various hospitals of Singapore. AKI-RiSc can then be sequentially applied to hospitals in regional South East Asian countries, and eventually to the international setting of the world abroad. We believe that the simplicity of AKI-RiSc and use of common clinical variables makes it a good candidate for validation in hospitals across different settings. Apart from assessing its predictive performance in a prospective validation cohort, it would also be essential to gain feedback from physicians on the score’s perceived utility and ease of use in helping them better care for their patients. AKI bundles can also be developed to complement the AKI-RiSc to assess its effectiveness in improving patient outcomes.

## Conclusions

In conclusion, we developed a simple and interpretable point-based AKI risk score which can be easily implemented on the ground for early identification of high-risk patients in the Emergency Department. It also has the potential be applied in healthcare settings internationally.

## Data Availability

Details of the variables and derived predictive model are available from the corresponding author.

## Supplementary Materials

**Figure S1:**
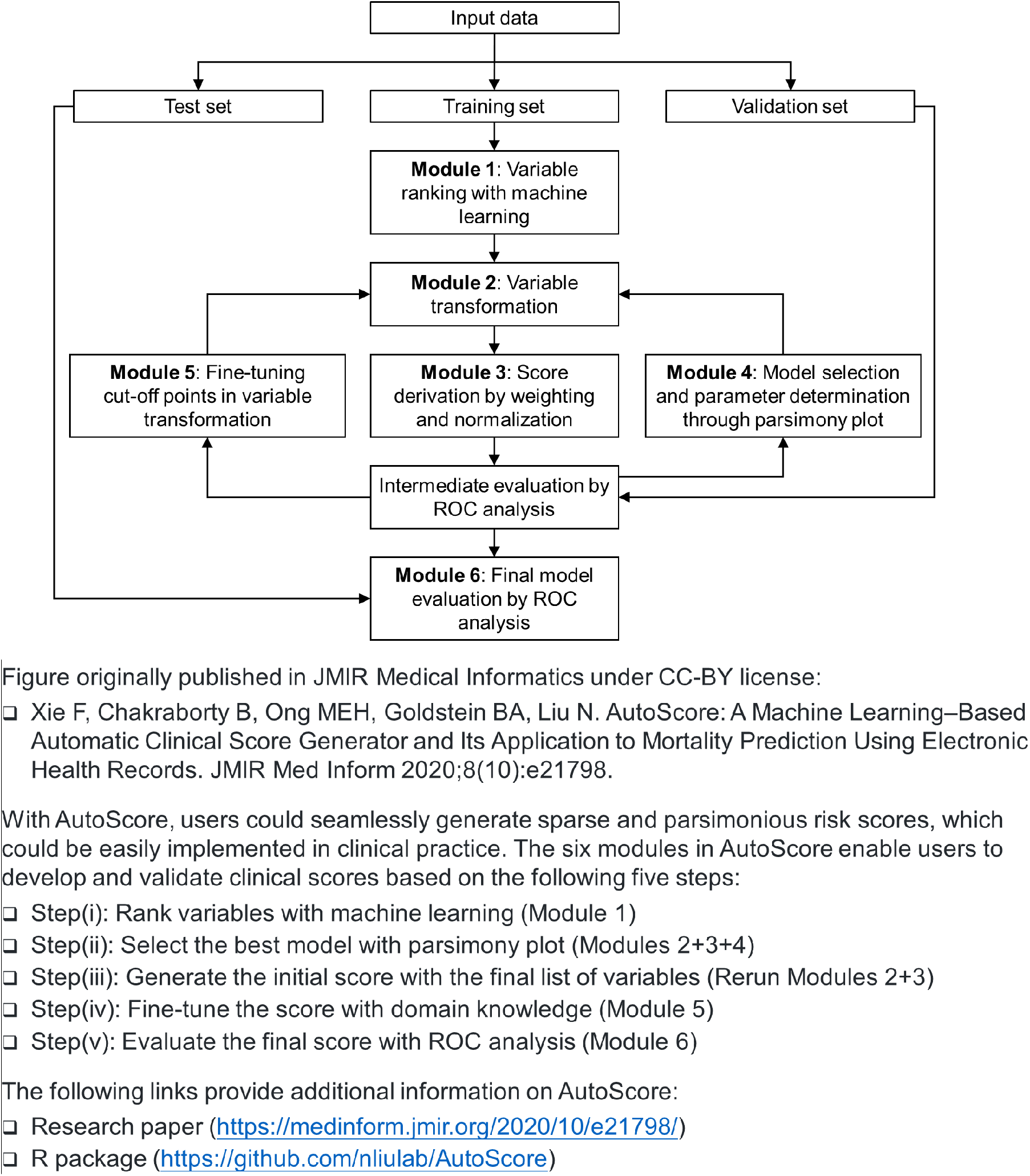
The AutoScore workflow.

## Notes

### Competing Interest Statement

The authors have declared no competing interest.

### Funding Statement

This research received funding from Duke-NUS Medical School.

### Author Declarations

A waiver of consent for EHR data collection was granted, and the study protocol was approved by Singapore Health Services' Centralised Institutional Review Board.

